# Towards Formal Computable Representation of Clinical Trial Eligibility Criteria for Alzheimer’s Disease

**DOI:** 10.1101/2022.03.21.22272707

**Authors:** Pengfei Yin, Hansi Zhang, Xing He, Matthew Diller, Qian Li, Shubo Tian, Arslan Erdenga-sileng, Zhe He, Cui Tao, Jiang Bian

**Author notes:** {, }, {, }.

## Abstract

Ambiguity and misunderstanding of free-text clinical trial eligibility can affect the accuracy of translating trial investigators’ mental model of the study population to the correct cohort identification queries. In this pilot study, to eliminate the ambiguity when parsing eligibility criteria, we built ontology-based representations to standardize clinical trial eligibility criteria. We analyzed 10 Alzheimer’s disease (AD) trials’ eligibility criteria and categorized them into general query patterns using an annotation schema borrowed from the literature on constructing knowledge graphs. Then, for each pattern, we built the corresponding ontological representations, linked them to real-word electronic health record (EHR) data, and constructed cohort identification queries using the neo4j graph database. Our evaluation results of these cohort queries verified the accuracy of our ontology representation; and interestingly, we found that graph-queries achieved better runtime performance for complex study traits. These results indicated that our approach is feasible and potentially beneficial; nevertheless, more systematic and comprehensive investigations are warranted.

## 1 Introduction

Eligibility criteria are critical in the clinical trials, which define the study population that shall participate in clinical trials (Chondrogiannis et al., 2017). Nevertheless, trial eligibility criteria are unstructured free-text written in natural language, where the ambiguity often causes researchers misunderstand the meaning of eligibility criteria and have difficulty parsing these criteria computationally into database queries (Tu et al., 2011). Misunderstanding of eligibility criteria and inaccurate cohort identification queries can lead to deviations from the original trial recruitment target, where (1) certain enrolled participants may not meet the requirements as the eligibility criteria described, or (2) missing potential eligible participants, which leads to low accrual of the trials. For example, an inclusion criterion “*creatinine < 1*.*5 times upper limits normal*” leads to ambiguity on what biospecimen source (e.g., serum, urine, blood) the creatinine is measured from.

Thus, standardized and computable knowledge representations for eligibility criteria are needed. Nevertheless, there is no consistent knowledge representation (e.g., standardized terminology, syntactic formats, and computability) for eligibility criteria (Weng et al., 2010). Ontology (i.e., a standardized representational framework that includes defined classes and relations among them (Arp et al., 2015) is a common tool for formal knowledge representations. There are a number of existing works on ontology-based representation of eligibility criteria. Milian et al. proposed an automated method, leveraging pattern detection and semantic tagging to turn free-text eligibility criteria into a structured format (Milian et al., 2015); nevertheless, it merely used UMLS concepts to standardize the entities rather than providing a computable representation of the criteria. Tao et al. created an ontology-based method focusing on representing temporal patterns in eligibility criteria (Crowe & Tao, n.d.); nevertheless, they did not fully consider all use cases (e.g., missing representation of the length of time an event occurs before the other event). Given the page limit, interested readers can refer to the review in (Weng et al., 2010) for a more comprehensive understanding.

In parallel, the literature on automatically building knowledge graphs with formal (often ontology-based) representations through mining free-text data (e.g., publications and web content) is rich. For example, a number of works has demonstrated the feasibility of using advanced natural language processing (NLP) methods to construct biomedical knowledge base/graphs in the format of triples (i.e., subject-predicate-object) to represent factual statements (e.g., SemRep (Rindflesch & Fiszman, 2003)). More recently, Jiang et al. proposed a novel scientific knowledge graph representation and construction model that considers not only the factual statements, but also the conditions when the factual statements are true (Jiang et al., 2019). In our past work on computable eligibility criteria (Zhang et al., n.d.), we have observed that conditions are frequently encountered and important when representing eligibility criteria. For example, in a criterion, “*Patient who had Parkinson’s disease within the prior 5 years,”* the temporal constraint *“within the prior 5 years”* is better represented as the condition of when the “*Parkinson’s disease*” diagnosis was given.

Thus, in this pilot study, we analyzed eligibility criteria from a small sample of Alzheimer’s disease (AD) trials and categorized them into general query patterns using an annotation schema borrowed from the literature on constructing knowledge graphs (Jiang et al., 2019). Then, we built formal ontology representations for the different query patterns, and further mapped them to electronic health records (EHRs) data from the OneFlorida Clinical Research Consortium in a graph database (i.e., Neo4j) to achieve semantic queries for cohort discoveries. To evaluate the accuracy of our translations, we compared our graph queries with manually curated data queries in Python pandas against the original data formats.

## 2 Methods

### 2.1 Data Sources

In this pilot, we randomly selected 10 Phase III/IV AD interventional trials and obtained their free-text eligibility criteria from *ClinicalTrials.gov*. We obtained individual-level patient EHR data from the OneFlorida Clinical Research Consortium – a clinical data research network contributing to the national Patient-Centered Clinical Research Networks (PCORnet). The OneFlorida data follows the PCORnet Common Data Model (CDM) that contains detailed patient and clinical variables, including demographics, diagnoses, procedures, vitals, medications, and labs. For performance reasons, we randomly selected 10% patient data from patients diagnosed with AD-related dementias (AD/ADRD) based on diagnostic (i.e., ICD-9/10) codes.

### 2.2 Define Patterns for Eligibility Criteria

We tailored an annotation guideline for eligibility criteria based on the study from Jiang et al. on knowledge graph construction (Jiang et al., 2019), where decomposed individual criteria into fact and condition tuples. Fact tuples are in the format of (subject, relation, object) which focus on the clinical observations (clinical event) related to a patient; and conditions represent the constraints related to the clinical observations such as temporal constraints of the clinical observations. Based on the annotation results, we summarized the criteria patterns using fact and condition tuples. The criteria patterns are categorized into demographic, diagnosis, encounter, medication, procedure, lab test respectively (PCORnet, 2020). Note that as we focused on developing computable representations, we excluded criteria that are not computable (e.g., patient willingness for consent) against the EHR data.

### 2.3 Ontology Construction and Representation of Eligibility Criteria

An ontology comprises defined classes and certain relations among them, which can help us standardize the entities and relations, but also potentially encode logics (e.g., temporal constraints) in the ontology. Based on our analysis of the eligibility criteria patterns, we created the initial version of the Computable Eligibility Criteria Ontology (CECO) using Basic Formal Ontology (BFO) as the upper-level ontology. We took a bottom-up approach (i.e., analyzing individual entities extracted from the eligibility criteria through the annotation process, defining the ontology classes corresponding to each entity, and establishing the relations among the ontology classes. Following best practice in ontology construction, we reused existing high-quality biomedical ontologies and iteratively improved the ontological structure. As the concept of computable eligibility criteria (against EHR data) is similar to computable phenotypes (i.e., “*clinical conditions, characteristics, or sets of clinical features that can be determined solely from EHRs and ancillary data sources*.”) (Tasker, 2017), we heavily reused the classes in the Human Phenotype Ontology (HP) (Köhler et al., 2017). Otherwise, we prioritized to reuse classes in existing well-known ontologies that follow the BFO as the upper-level ontology and the scope of the ontology is similar to CECO (e.g., (Arp et al., 2015)). For the temporal constraints, we reused the Time Event Ontology (TEO) to represent events, time points, and their relationships.

### 2.4 Query Construction and Evaluation

We imported the CECO ontology into Neo4j a graph database platform (Neo4j, 2021) using the neosemantics toolkit (Neo4j Lab, 2021). According to the representations in CECO, we represented the criteria patterns as (knowledge) graphs in Neo4j. We then imported the individual-level AD/ADRD patient data from OneFlorida and linked classes of CECO to the data elements in OneFlorida. We evaluated the accuracy of the our representations by comparing the graph query results with the results from directly applying query logic (i.e., Python and pandas) to the RWD.

## 3 Results

### 3.1 Patterns for Alzheimer’s disease (AD) Trials

From the 10 AD trials, we identified 157 eligibility criteria, where each trial has an average of 4.7 (2 - 9) inclusion and 11 (3 - 25) exclusion criteria. We further decomposed each criterion into study traits (i.e., the minimal units that do not change the meaning of the original criterion; e.g., the inclusion criterion “*patients with AD but not with other AD-related dementia*” can be decomposed into two study traits (1) “*patients with AD*”; and (2) “*patients without other AD-related dementia*”). In sum, we identified 223 computable (and ignored those that are not computable as our focus is representation for data queries) study traits (i.e., merged inclusion and exclusion study traits, as (1) the relationship between the two is simply negation, and (2) a study trait can be either inclusion or exclusion depending on the specific trials), which would not affect how the we summarize the patterns of criteria/study traits patterns. Through annotation, we identified and formalized 9 types of criteria patterns based on data domains as shown in **Table 1**. For example, a criterion “*Patient who had Parkinson’s disease within the prior 5 years*.” can be classified as a diagnosis pattern and represented as follows:

**Table 1.** Identified criteria patterns from the 10 AD trials.

| Table 1. Identified criteria patterns from the 10 AD trials. |  |  |
| --- | --- | --- |
| Type | Fact/Cond. | Tuples |
| **Demographic (Dem.)** | Fact | ({subject: age}, greater than or equal to, number) |
|  | Fact | ({subject: sex}, equals to, xxx) |
| **Encounter** | Fact | (subject, has encounter status, encounter type) |
| **Procedure** | Fact | (subject, treated with, procedure) |
| **Diagnosis (DX)** | Fact 1 | (subject, diagnosed with/not diagnosed with, disease) |
|  | Fact 2 | (subject, status, specific status) |
|  | Cond. 1 | (disease, {within/at least before: T time}, event) |
| **Medication (Med.)** | Fact | (subject, treated with/not treated with/are being treated with, drug) |
|  | Cond. 1 | (drug, {within/at least before: T time}, event) |
|  | Cond. 2 | (drug, less/greater/equal, {dosage: value}) |
|  | Cond. 3 | (drug, more than, one type) |
| **Lab test** | Fact | (subject, has lab test result, lab test result) |
|  | Cond. 1 | (lab test result, {within/at least before: T time}, event) |
|  | Cond. 2 | (lab test result, greater/less/equal, {result: value}) |
| **DX + Encounter** | Fact | (subject, diagnosed with/not diagnosed with, disease) |
|  | Cond. 1 | (disease, {within before: T time}, event) |
|  | Cond. 2 | (subject, has encounter status, encounter type) |
|  | Cond. 3 | (subject, has been treated for, disease) |
| **DX + Med.** | Fact | (subject, status, specific status) |
|  | Cond. | (subject, treated with, drug) |
| **DX + Dem.** | Fact | ({subject: sex}, diagnosed with, disease) |
|  | Cond. | (disease, {at least before: T time}, event) |

<sup>a</sup> The Fact/Cond. classification is verbatim of how they were classified in the limited number of eligibility criteria we reviewed; and a fact tuple in one criterion can be a condition tuple in another criterion.
<sup>b</sup> Note that certain criterion covers multiple data domains.

- Fact: (subject, diagnosed with, Parkinson’s disease)
- Condition: (Parkinson’s disease, {within T time before: 5 years}, baseline)

As another example, for criterion “*if subjects are taking Acetylcholinesterase inhibitors (AChEIs), they must be on a stable dose for > 3 months prior to baseline*,” it can be summarized as a medication pattern as:

- Fact: (subject, treated with stable dosage, AChEIs)
- Condition: (AChEIs, {before at least T time: 3 months}, screening visit)

Note that a fact tuple in one criterion can be a condition tuple depending on the individual criteria; and a complex criterion would use a mix of the different patterns with different logic operations.

The other important aspect of modeling criteria is temporal patterns, as much of a criterion expresses how different events are related to each other on the patient timeline. As in our previous example, i.e., “*(Parkinson’s disease, {within T time before: 5 years}, baseline)*,” it describes that the patient has to have been diagnosed with Parkinson’s disease (i.e., event X) within 5 years (i.e., a temporal constraint) before the baseline (i.e., when the screening happens as event Y) as shown in Figure 1. We have summarized temporal patterns previously in (Zhang et al., n.d.).

**Figure 1.**
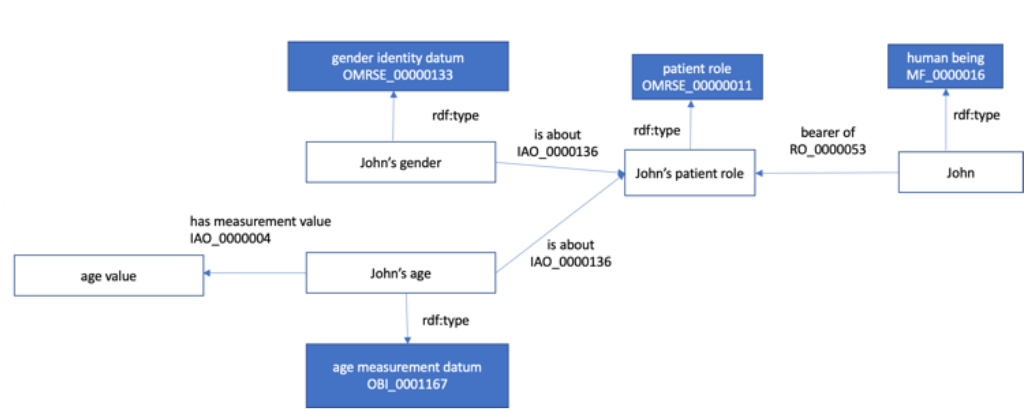
The ontology representation of patient demographics.

### 3.2 Ontology Representation

We used CECO classes and relations to standardize and represent the eligibility criteria patterns we discovered.

#### Demographic

The first step is to represent patients, where we consider patient as role of human being (dependent relationship). We used “*bear of*” relation from relation ontology (RO) to describe the dependent relationship where the existence of patient roles depends on the existence of human being. To represent patient demographic information such as age, we used “*is about*” relation to categorize these attributes as information artifact of a human being. We introduce the data property “*has measurement value*” to capture the age value of the patients. An example of the demographic representation is shown in Figure 2.

**Figure 2.**
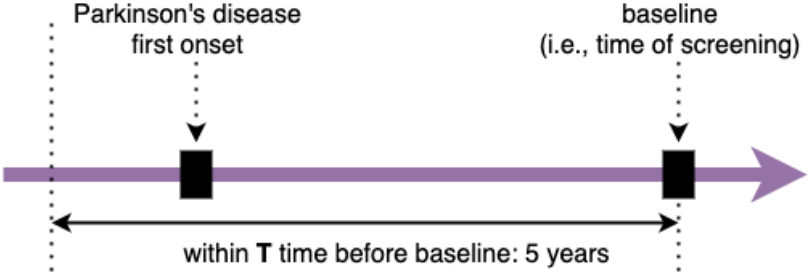
An example of temporal constraint for the criterion, *“(Parkinson’s disease, {within T time before: 5 years}, baseline).”*

#### Encounter

Encounter is a subclass of the health care process. We used “*has participant*” relationship from RO to represent patients (i.e., human being with patient role) that participate in the process of health care encounters. For temporal relations, we used object property “*hasValidTime*” in TEO ontology, where we consider clinical observations as events and describe the temporal constraint by linking an clinical event (individual) with a specific time (individual) (Li et al., 2020) as shown in Figure 3.

**Figure 3.**
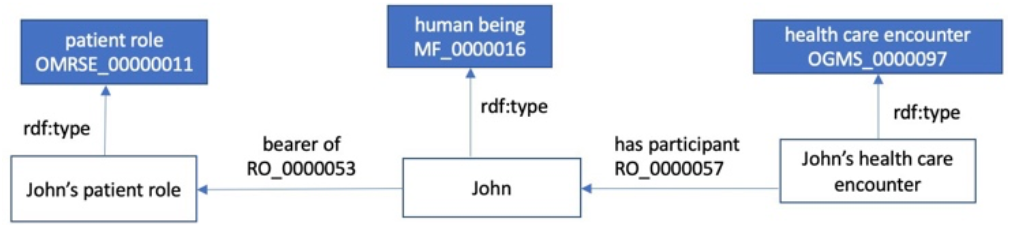
The ontology representation of a patient encounter.

#### Procedure

We only found one criterion related to procedures in our limited criteria sample, which simply described a subject treated with a type of procedure. We can find the “*medical procedure*” in the Ontology of Medically Related Social Entities (OAE), which is a subclass of “*medical intervention*”; nevertheless, “*medical intervention*” also contains other treatment related procedures (e.g., “*acupuncture*” as we found in one of criteria we reviewed). For simplicity, we used “*medical intervention*” in Figure 4 to demonstrate these procedure patterns. Further, the procedure would only happen through the patient’s encounter with the health care system, as shown in Figure 4.

**Figure 4.**
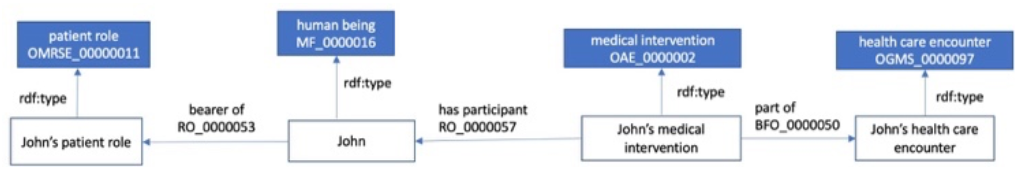
The representation for procedures.

which is the bearer of the “*specimen role;”* and patients participated in the “*specimen collection process*” for the collection of the specimen. A clinical laboratory test is performed on the specimen. Thus, the specimen is an input of the lab test, and the lab result is the laboratory finding of the lab test.

#### Diagnosis

The representation of diagnosis is based on Hogan *et al*. work (Hogan & Ceusters, 2016). Further, a diagnosis of a certain disease does not necessarily mean the patient has that disease, but can be an artifact of the diagnostic process. Thus, we represented these two separately: (1) a diagnosis given as part of a health care encounter (i.e., Fact 1 in DX type); and (2) a disease as a phenotype abnormality that a patient has (i.e., inheres in; Fact 2 in DX type). For temporal relations, we considered the diagnostic process as an event since the events are time-oriented (Li et al., 2020). The diagnosis representation is shown in Figure 5.

**Figure 5.**
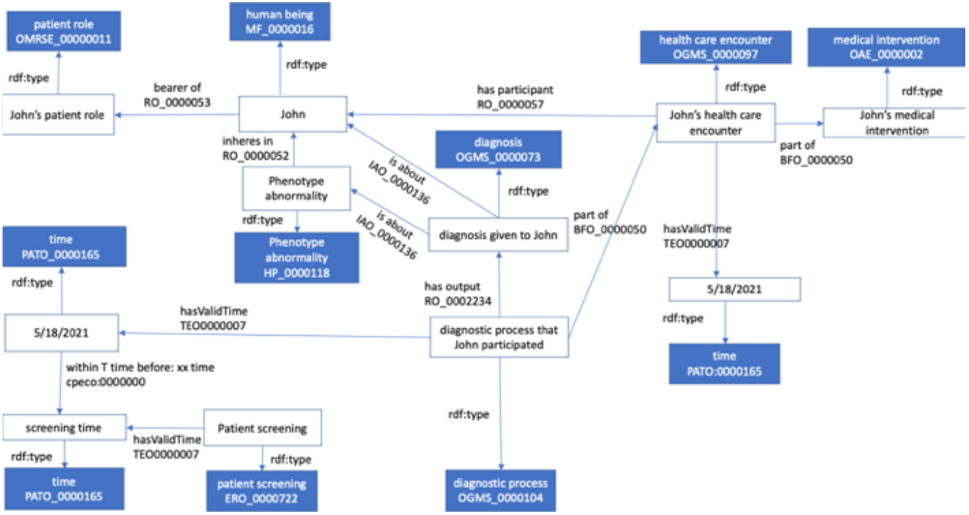
The representation of diagnosis.

#### Medication

As shown in Figure 6, we consider the “*drug administration*” as a process; the patient role is “*realized in*” the “*drug administration*” process; and the “*drug product*” is the participant of the “*drug administration*” process. To capture temporal constraints, we can consider the “*drug administration*” process as an event. We used the class “*dose*” from the Ontology for Biomedical Investigations (OBI) and linked it with the “*drug product*” using “*is about*” relation, where we described the drug dosage as an information entity about the “*drug product*”.

**Figure 6.**
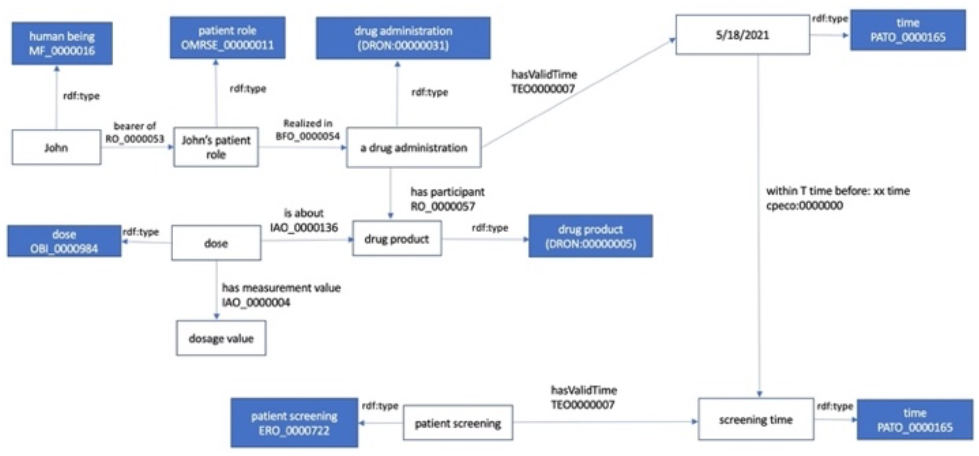
The representation of medications.

#### Lab test

The representation of lab tests and results is shown in Figure 7. We defined the “*specimen”* as the material entity

**Figure 7.**
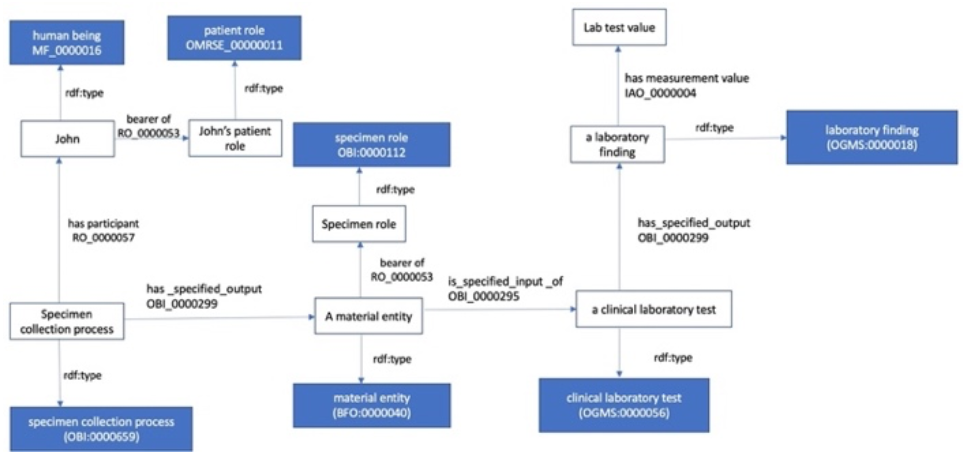
The representation of lab tests.

### 3.3 Query Construction and Evaluation

Built upon these representations, we constrctured graph-based semantic queries against the RDF data in neo4j for a range of randomly sampled study traits and compared the query results with the queries constructred in Python pandas against the original datasets (i.e., relational database following the PCORnet CDM converted into csv files). All graph-based queries returned the same results as the pandas-based queries indicating query and representation accuracies. We also recorded the runtimes of each graph-query and comparied with their pandas conterparts. Interestingly, for complex study traits (e.g., “*patients has been hospitalized or treated for suicidal behavior in the past 5 years*”), graph queries are faster than pandas queries (e.g., 0.106 seconds vs. 0.2452 seconds, respectively, for this example); while for simple study traits (e.g., “*aged ≥ 50 years*”), graph queries are slower than their pandas conterparts (e.g., 0.021 seconds vs. 0.00198 seconds, respectively, for this example). Nevertheless, our sample size is small, and more systematic and comprehensive evaluation is needed.

## 4 Discussion and Conclusion

We summarized eligibility criteria patterns of 10 AD trials and created the CECO to standardize and render them computable through graph-based queries using neo4j graph database. Our study is limited as we only analyze 10 AD trials; thus, the identified patterns are still unrepresentative. Nevertheless, we initial evaluations are satisfactory in terms of both the accuracy of the ontological representation and graph-query results. Interestingly, we found that graph-based queries outperform their traditional counterparts (i.e., pandas against relational csv files in this pilot study), suggesting potential benefits of using a graph-based ontological approach besides having a more accurate mental model to render the eligibility criteria computable. Thus, a more systematic and comprehensive study is warranted.

## Data Availability

All data produced in the present study are available upon reasonable request to the authors

## Acknowledgments

This study is funded in part by the National Insulites of Health (NIH) through awards: R21 AG068717 and R21 CA253394.

